# Substance use in youth at genetic and clinical high risk for psychosis

**DOI:** 10.1101/2022.12.01.22282991

**Authors:** Carolyn M. Amir, Simon Kapler, Gil Hoftman, Leila Kushan, Jamie Zinberg, Kristin S. Cadenhead, Leda Kennedy, Barbara A. Cornblatt, Matcheri Keshavan, Daniel H. Mathalon, Diana O. Perkins, William Stone, Ming T. Tsuang, Elaine F. Walker, Scott W. Woods, Tyrone D. Cannon, Jean Addington, Carrie E Bearden

## Abstract

**Background:** Elevated rates of alcohol, tobacco, and cannabis use are observed in both patients with psychotic disorders and individuals at clinical high risk for psychosis (CHR-P), and strong genetic associations exist between substance use disorders and schizophrenia. While individuals with 22q11.2 deletion syndrome (22qDel) are at increased genetic risk for psychosis, initial evidence suggests that they have strikingly low rates of substance use. In the current study, we aimed to directly compare substance use patterns and their neurobehavioral correlates in genetic and clinical high-risk cohorts.

**Methods:** Data on substance use frequency and severity, clinical symptoms and neurobehavioral measures were collected at baseline and at 12-month follow-up visits in two prospective longitudinal cohorts: participants included 89 22qDel carriers and 65 age and sex-matched typically developing (TD) controls (40.67% male, M_age_=19.26 ± 7.84 years) and 1288 CHR-P youth and 371 matched TD controls from the North American Prodrome Longitudinal Study-2 and 3 (55.74% male; M_age_=18.71 ± 4.27 years). Data were analyzed both cross-sectionally and longitudinally using linear mixed models.

**Results:** Controlling for age, sex, and site, CHR-P individuals had significantly elevated rates of tobacco, alcohol, and cannabis use relative to TD controls, whereas 22qDel had significantly lower rates. Increased substance use frequency and severity in CHR-P individuals was associated with increased positive psychosis symptom severity, dysphoric mood, social functioning, and IQ, while higher social anhedonia was associated with lower substance use frequency and severity, across all domains at baseline. These patterns persisted when we investigated these relationships longitudinally over one-year. CHR-P youth exhibited significantly increased positive psychosis symptoms, dysphoric mood, social anhedonia, and IQ compared to 22qDel carriers, and significantly higher social functioning and lower rates of autism spectrum disorder (ASD) compared to 22qDel carriers, both at baseline and at one year follow-up.

**Conclusions:** Individuals at genetic and clinical high risk for psychosis have strikingly different patterns of substance use. Factors such as increased neurodevelopmental symptoms (lower IQ, higher rates of ASD) and poorer social functioning in 22qDel may help explain this distinction from substance use patterns observed in CHR-P individuals.

## Introduction

Excessive substance use occurs at elevated rates in patients with psychotic disorders (Blanchard et al., 2000; Alvarez-Jimenez et al., 2012; Addington et al., 2014; Khokhar et al., 2018; Ward et al., 2018; Archibald 2019; Catthor & Dom, 2022; Correll et al., 2022) and cannabis use in particular has been suggested to play a role in the onset of psychosis (Arseneault et al., 2002; Caspi et al., 2005; Moore et al. 2007; Foti et al., 2010; Kuepper et al. 2011; Marconi et al., 2016; Mustonen et al., 2018; Hasin et al., 2020; Hasan et al., 2020). Patients with schizophrenia are 4.6 times more likely to use and abuse substances than the general population (Refier et al. 1990). Similar rates of hazardous substance use, ranging from 22% to over 50%, are reported for individuals at clinical high risk for psychosis (CHR-P)— individuals experiencing attenuated psychotic-like experiences preceding the onset of psychosis (Addington et al., 2014; van der Meer et al., 2012)—compared to typically developing controls (TD; Santesteban-Echarri et al., 2022; Stevenson et al., 2022; Farris et al., 2020; Buchy et al. 2015; Addington 2014). In contrast, a notable lack of substance use has been reported in people with chromosome 22q11.2 deletion syndrome (22qDel; Vingerhoets et al. 2019; Basset et al. 2003), one of the strongest genetic risk factors for psychosis (Rees et al. 2017; Marshall et al. 2017; Malhorta et al. 2012; Green et al. 2009; Chow et al. 2006).

The reasons for the substantially elevated rates of hazardous substance use in individuals with idiopathic psychotic disorders are not fully understood but are thought to be linked to the brain’s dopaminergic pathways, associated with the reward properties of drugs and positive symptoms of psychosis (Khokar 2018; Volkow 2009). Certain genetic factors lead to an increased risk of substance use and psychotic disorders, and genetic risk is thought to interact with environmental factors including the social environment (Moffitt et al., 2005; Fanous & Kendler, 2008; Henquet et al., 2008; Khokhar et al. 2018; Gillespie & Kendler, 2021). That individuals with 22qDel may need little or no environmental insult to develop psychosis is consistent with the liability threshold model; stronger genetic predisposition may ‘tip the scales’ towards phenotypic expression of psychosis, even in the absence of environmental risk factors (McGue et al., 1983, Tsuang et al., 2001, McGuffin, 2004; Martin et al. 2014).

While not yet elucidated in 22qDel, neurobehavioral traits associated with hazardous substance use have been studied in CHR-P. In particular, positive symptom severity is greater in CHR-P who use illicit substances compared to CHR-P non-users and ultra high risk individuals have higher rates of unusual thought content and suspiciousness compared to ultra high risk non-users (Carney et al., 2017; van der Meer et al., 2012; Schifani et al., 2020). Relative to CHR-P without cannabis use, CHR cannabis users have significantly lower levels of social anhedonia (Gill et al., 2015; Kolliakou et al., 2015; Santesteban-Echarri et al., 2022). A proposed explanation for this inverse association is that CHR-P with lower levels of social anhedonia may tend to engage more with peers and seek substance use for social motives (Santesteban-Echarri et al., 2022). Further, CHR-P and individuals with first-episode psychosis report social engagement as a primary reason for cannabis use (Gill 2015; Gomez Perez 2017; Kolliakou 2015) and higher levels of social functioning are associated with elevated substance use rates in CHR-P (Author et al., 2012). While not yet studied in CHR-P, higher peer engagement has been found to be associated with increased risk for substance use particularly during adolescence (Keefe 1994; Kuther & Higgins-D’Alessandro 2003; Whitesell et al., 2013; Brooks-Russel et al. 2014), which has been theorized to be attributable to an increase in substance use accessibility with higher social engagement. Mood enhancement is also cited by CHR-P as a primary motivation for substance use seeking (Gill 2015), and while the association between mood and substance use has not yet studied in CHR-P, greater depression symptoms are associated with elevated substance use rates in adolescence (Rao 2006). CHR-P cannabis lifetime users have higher intelligence quotient (IQ) scores compared to non-users, which may be attributable to higher premorbid cognitive function (Ferraro et al., 2019).

To our knowledge, this is the first study examining substance use in CHR-P and 22qDel in parallel which offers a unique opportunity to contrast clinical and genetic factors underlying hazardous substance use during adolescent development. Investigating factors associated with decreased rates of substance use in 22qDel could point to a protective phenomenon, with differential implications for prediction and treatment/prevention in subsets of individuals at high risk for psychosis. Here, we investigated rates of substance use in two prospective longitudinal cohorts, youth with CHR-P symptoms and youth with 22q11.2 deletions, as well as demographically-matched typically developing (TD) controls, with the hypothesis that CHR-P youth will have elevated rates of substance use relative to TD controls, both cross-sectionally and longitudinally over one year, whereas 22qDel participants will not. Second, we aimed to investigate the relationship of substance use to psychiatric symptoms and neurobehavioral traits. We predicted that better social functioning and higher IQ, as well as increased dysphoric mood and psychosis symptom severity would be associated with increased rates of substance use, while social anhedonia would be inversely related with substance use. We then examined differences in these measures between 22qDel carriers and CHR-P youth to elucidate the potential influence of social functioning, social anhedonia, dysphoric mood, psychosis symptom severity, and IQ on substance use patterns in this population.

## Methods

### Participants

This study examined substance use over time in both individuals with molecularly confirmed 22q11.2 deletions (n = 89 22qDel, 45% male) and age and sex-matched typically developing controls (TD; n = 65, 46.15% male) and in CHR-P youth and age and sex-matched typically developing controls. CHR-P participants were recruited as part of the North American Prodromal Longitudinal Studies 2 (NAPLS 2; Addington 2012) and NAPLS 3 (Addington et al., 2022), designed to investigate predictors and mechanisms of transition to overt psychosis. This paper reports on 737 CHR-P youth (57.53% male) and 275 TD control subjects (50.18% male) who completed substance use assessments. Use of the NAPLS 3 cohort in this study is intended to replicate our NAPLS 2 baseline findings in a population at higher risk for conversion to psychosis (at “enhanced” risk; 40% likelihood of psychosis conversion; Addington et al., 2022; Cannon et al., 2016) to test whether findings hold in an enhanced sample. NAPLS 3 participants include 551 “enhanced” CHR-P participants (56.99% male) and 96 TD control subjects (50% male). Detailed descriptions of the recruitment procedures and measures are reported elsewhere (Addington et al. 2012; Addington et al., 2022).

Individuals with 22qDel included in this study represent a subset of participants ascertained as part of an ongoing longitudinal study at the University of California, Los Angeles. Details of the recruitment and methods for this study are described elsewhere (Lin et al., 2020). Age and sex-matched TD participants were recruited from local communities via web-based advertisements and flyers/brochures in local schools, pediatric clinics, and other community sites. Exclusion criteria for all study participants included significant neurological or medical conditions (unrelated to 22qDel) that might affect brain structure or function, history of head injury with loss of consciousness, and insufficient fluency in English.

CHR individuals and TD controls were recruited for NAPLS studies, which were approved by the Institutional Review Boards of all eight NAPLS sites. Written informed consent, including parental consent, was obtained from all adult participants and parents/guardians of minors. The University of California, Los Angeles Institutional Review Board approved all study procedures and informed consent documents related to 22q11.2 CNV studies. Participants under the age of 18 years provided written assent, while their parent or guardian completed written consent in each study. Across both cohorts, we restricted our analyses to participants age 12 and older. As maximal data were available for the first two timepoints of the studies, longitudinal analyses included baseline visits and one-year follow-up visits.

### Clinical Status

A summary of measures and timepoints analyzed in this study are presented in Table 1. The SIPS and the Scale for Assessment of Psychosis-Risk Syndromes (SOPS; McGlashan et al. 2010) were used to define CHR-P, rate dimensional psychosis-risk symptom severity and development of overt psychosis in both cohorts. Individual items were rated, and summary scores were determined for each domain (positive, negative, disorganized, and general symptoms). Total positive symptoms were defined as the sum of items in the positive symptom domain. Dysphoric mood was assessed via an item on the general symptom subscale, and social anhedonia as an item on the negative symptoms subscale. Full details regarding SIPS criteria, reliability and consensus procedures are described elsewhere (Addington et al. 2012; Addington et al., 2022). The SCID was used to establish *DSM-IV/V* Axis I diagnoses at baseline and follow-up for all participants, including ASD diagnosis. Clinical raters in both studies were experienced research clinicians; ratings and diagnoses were additionally validated via case consensus procedures (Addington et al., 2012; Addington et al., 2022; Lin et al., 2019). IQ in CHR-P was assessed by the Wechsler Abbreviated Scale of Intelligence (Wechsler 1999) and in 22qDel carriers was assessed by the Full-Scale Intelligence Quotient.

**Table 1.** Overview of assessments by sample and timepoint

| Assessment | NAPLS 2 baseline | NAPLS 2 12 months | NAPLS 3 baseline | NAPLS 3 12 months | 22qDel baseline | 22qDel 12 months |
| --- | --- | --- | --- | --- | --- | --- |
| Clinical Diagnosis* | SCID | SCID | SCID | SCID | SCID | SCID |
| Psychosis symptoms | SIPS/SCID | SIPS/SCID | SIPS/SCID | SIPS/SCID | SIPS/SCID | SIPS/SCID |
| Alcohol/drug use | AUS/DUS | AUS/DUS | AUS/DUS | - | SCID + CBCL | SCID + CBCL |
| Social functioning | Global functioning scale: Social | Global functioning scale: Social | Global functioning scale: Social | Global functioning scale: Social | Global functioning scale: Social | Global functioning scale: Social |
| Intelligence quotient (IQ) | WASI-2 FSIQ | WASI-2 FSIQ | WASI-2 FSIQ | WASI-2 FSIQ | WASI-2 FSIQ | WASI-2 FSIQ |
| Autism Spectrum Disorder | SCID | -- | SCID | -- | SCID/ADI/ADOS | -- |

*\*Modified SCID used across 22qDel and NAPLS studies includes an additional developmental disorders module (Foss-Feig et al., 2019)*

### Substance Use

Substance use in NAPLS 2 and NAPLS 3 participants was assessed using the Alcohol Use Scale/Drug Use Scale (AUS/DUS; Drake et al. 1996) at all visits. Both severity (1 = abstinent, 2 = use without impairment, 3 = abuse, 4 = dependence) and frequency of substance use (0 = no use, 1 = once or twice per month, 2 = 3–4 times per month, 3 = 1–2 times per week, 4 = 3–4 times per week, 5 = almost daily) were collected for tobacco, alcohol, marijuana, cocaine, opiates, phencyclidine (PCP), amphetamines, methylenedioxy-methylamphetamine (MDMA), γ-hydroxybutyric acid (GHB), huffing (inhaling glue, other volatiles), hallucinogens, and other drugs. Frequency of tobacco use was the only item rated differently (0 = no use, 1 = occasionally, 2 = less than 10 times per day, 3 = 11–25 per day, 4 = more than 25 per day). Longitudinal group comparisons were conducted for NAPLS 2 data; baseline results were analyzed to test for replication in NAPLS 3, as control participants did not complete the AUS/DUS at follow-up in NAPLS 3. 22qDel and respective TD control participants were assessed at each timepoint via SCID interview for substance use/abuse within the past 6 months, and the Child Behavior Checklist (CBCL; Achenbach et al. 1991) was used to assess lifetime substance use or abuse, including alcohol, non-prescription drug use, and tobacco use. The CBCL was completed by parents of participants aged 12-18.

### Social Functioning

The Global Functioning: Social scale was used to measure social functioning in both cohorts. Social functioning is calculated as highest level of social functioning in the past year. Developmentally appropriate, detailed descriptions are provided to illustrate the range of functioning captured by each point on the scale, with lower scores indicating more impairment (Auther et al. 2006).

#### Analyses

Analyses were conducted in R 4.1.2 using statistical software package lme4 (Bates et al. 2015). We compared individuals with 22qDel to respective age and sex-matched TD controls, and CHR-P youth to a separate age and sex-matched TD control group (with TD controls as the reference groups). For NAPLS-2 and 22qDel cohorts, timepoints included intake visit and one-year follow-up visits, and longitudinal models included a group-by-time interaction. The use of substances other than alcohol, tobacco or cannabis was either minimal or absent in all samples; therefore, only alcohol, tobacco, and cannabis were considered for further statistical analysis. To determine whether CHR-P and 22qDel show different rates of substance use relative to TD controls, linear mixed models with substance use as the dependent variable and participant group as the independent variable were used to test for group differences in substance use (with controls as the reference group). We tested replication of NAPLS 2 results with NAPLS 3 baseline data, as substance use information was not collected longitudinally in NAPLS 3. Age, sex, and site were included covariates, and participant ID was included as the random effects term. FDR correction was applied within each model applied on effects of subject group.

To test relationships between substance use and clinical symptoms, linear mixed models with each symptom domain as dependent variable (DV) and substance use characterization as independent variable (IV) were tested for control and CHR-P participants at baseline in NAPLS 2 and NAPLS 3. In NAPLS 2, we tested relationships between baseline clinical symptoms and substance use at follow-up in CHR-P, additionally controlling for baseline substance use rates. Main effects of each model were FDR-corrected for multiple comparisons.

We then tested differences in clinical symptom measures between 22qDel participants and CHR-P to elucidate neurobehavioral traits influencing substance use patterns in this population (with 22qDel carriers as the reference group). Linear mixed models with substance use as the DV and participant group as the IV were used to test for differences in clinical symptom measures at baseline and longitudinally with participant age, sex, and site included as covariates. FDR correction was applied within each model applied on effects of subject group.

## Results

### Descriptives

Demographic variables at baseline are presented for each group in Table 2 and Table 3. There were significant differences in medication use between 22qDel carriers and TD controls, as well as between CHR-P and TD controls, such that TD controls had lower rates of medication use. In the CHR-P group there were significantly higher rates of Hispanic TD control participants compared to the 22qDel group and significantly higher rates of non-white participants.

**Table 2:** Baseline characteristics: 22qDel carriers and TD Controls

| Baseline | Typically Developing Control Subjects | 22q11.2 Deletion Carriers |
| --- | --- | --- |
| N, total | 61 | 89 |
| Age, Years (SD), Age range<sup>b</sup> | 17.18 (4.28); 12-28 | 20.17(10.18); 12-61 |
| Males, n (%) | 29(47.5%) | 40(45%) |
| Non-white, n (%)<sup>a</sup> | 22(39.3%) | 8 (8.99%) |
| Hispanic<sup>a</sup> | 17 (27.87%) | 14 (15.73%) |
| Psychosis n (%)<sup>b</sup> | 0(0.00%) | 12(13.48%) |
| ASD, n (%)<sup>b</sup> | 0(0.00%) | 37(41.57%) |
| Alcohol use (%)<sup>a</sup> | 10(16.39%) | 3(3.37%) |
| Substance use (non-prescription)<br>(%) | 2(3.28%) | 2(2.24%) |
| Tobacco use | 2(3.28%) | 1(1.12%) |
| Medication, n (%)<sup>b</sup> | 2(3.28%) | 41(46.07%) |
| Anti-Psychotics | 0(0%) | 15(16.85%) |
| Mood Stabilizers | 0(0%) | 13(14.6%) |
| Stimulants | 1(1.54%) | 5(5.62%) |
| Other Medication | 1(1.54%) | 8(9.00%) |
| No Medication | 57(93.44%) | 48(53.93%) |

<sup>a</sup>Control > 22qDel ( $p < .05$ )
<sup>b</sup>22qDel > Control ( $p < .05$ )

**Table 3.** Baseline Characteristics: NAPLS 2 & 3 (CHR-P Subjects and TD Controls)

| **NAPLS 2 Baseline** | **TD Controls** | **CHR-P** |
| --- | --- | --- |
| N, total | 275 | 737 |
| Age, Years (SD), Age range<sup>a</sup> | 19.76(4.68); 12-34 | 18.49(4.24); 12-35 |
| Males, n (%) | 138(50.18%) | 424(57.53%) |
| Non-white, n (%) | 125 (45.45%) | 310(42.06%) |
| Hispanic | 48 (17.45%) | 138 (18.72%) |
| Medication, n (%)<sup>a</sup> | 7(2.55%) | 296(40.16%) |
| Anti-Psychotics | 0(0%) | 132(17.91%) |
| Mood Stabilizers | 0(0%) | 26(3.53%) |
| Stimulants | 3(1.09%) | 52(7.06%) |
| Other Medication | 4(1.45%) | 86(11.67%) |
| No Medication | 262(95.27%) | 441(59.84%) |
| **NAPLS 3 Baseline** | TD Controls | Clinical High Risk for Psychosis - Enhanced Subjects |
| N, total | 96 | 551 |
| Age, Years (SD), Age range | 18.60(4.22); 12-30 | 18.42(4.04); 12-30 |
| Males, n (%) | 48 (50%) | 314(56.99%) |
| Non-white, n (%)<sup>b</sup> | 51(53.13%) | 244(44.28%) |
| Hispanic | 24 (24.00%) | 125 (22.69%) |
| Medication, n (%)<sup>a</sup> | 1(1.04%) | 256 (46.46%) |
| Anti-Psychotics | 0(0.0%) | 114 (20.69%) |
| Mood Stabilizers | 0(0.0%) | 21 (3.81%) |
| Stimulants | 1(1.04%) | 40 (7.26%) |
| Other Medication | 0(0.0%) | 81 (14.7%) |
| No Medication | 0(0.0%) | 294 (53.36%) |
| <sup>a</sup>CHR-P > Control ($p<.05$) | | |
| <sup>b</sup>Control > CHR-P($p<.05$) | | |

### Substance use at baseline and over time

Results of mixed models revealed that CHR-P had greater frequency and severity of both cannabis and tobacco use compared with controls both at baseline and longitudinally (Table 4). There was no statistical difference in alcohol use severity between controls and CHR-P (see Tables 4). All group differences in baseline substance use observed in NAPLS-2 replicated in NAPLS 3 (see Table S2). Controlling for tobacco use in cannabis models and psychotropic medication use in all substance use models did not affect results.

**Table 4.** Differences in Substance Use between CHR-P and TD control Groups in NAPLS 2

|  | Effect of Subject Group |  | Effect of Time |  | Group * Time Interaction |  |
| --- | --- | --- | --- | --- | --- | --- |
| | $\beta$ | q-value | $\beta$ | p-value | $\beta$ | p-value |
| **Cross-sectional** |  |  |  |  |  |  |
| Alcohol Use Frequency | -.052 | .312 | - | - | - | - |
| Alcohol Use Severity | -.027 | .421 | - | - | - | - |
| Cannabis Use Frequency | **.348** | **<.001** | - | - | - | - |
| Cannabis Use Severity | **.384** | **<.001** | - | - | - | - |
| Tobacco Use Frequency | **.424** | **<.001** | - | - | - | - |
| Tobacco Use Severity | **.437** | **<.001** | - | - | - | - |
| SIAS | **1.23** | **<.001** | - | - | - | - |
| GFS highest | **-.539** | **<.001** |  |  |  |  |
| SAS | **1.31** | **<.001** | - | - | - | - |
| Positive Symptoms (SIPS) | **1.869** | **<.001** | - | - | - | - |
| Dysphoric Mood | **1.518** | **<.001** | - | - | - | - |
| Social Anhedonia | **1.21** | **<.001** | - | - | - | - |
| **Longitudinal** |  |  |  |  |  |  |
| Alcohol Use Frequency | -.01 | .757 | **.079** | **.006** | -.034 | .393 |
| Alcohol Use Severity | .012 | .752 | **.09** | **.004** | -.057 | .184 |
| Cannabis Use Frequency | **.160** | **<.001** | .055 | .061 | -.026 | .532 |
| Cannabis Use Severity | **.174** | **<.001** | .051 | .112 | -.027 | .538 |
| Tobacco Use Frequency | **.375** | **<.001** | .029 | .263 | -.035 | .331 |
| Tobacco Use Severity | **.207** | **<.001** | .029 | .26 | -.036 | .313 |
| SIAS | **.632** | **<.001** | -.039 | .131 | **-.15** | **<.001** |
| GFS highest | **-.580** | **<.001** | .010 | .668 | .055 | -.78 |
| SAS | **.718** | **<.001** | -.027 | .303 | **-.242** | **<.001** |
| Positive Symptoms | **.994** | **<.001** | -.027 | .192 | **-.353** | **<.001** |
| Dysphoric Mood | **.818** | **<.001** | -.023 | .408 | **-.295** | **<.001** |
| Social Anhedonia | **.629** | **<.001** | -.016 | .522 | **-.148** | **<.001** |

**Table S1.** Baseline substance use in NAPLS 2 and 3

| NAPLS 2 | Alcohol |  | Tobacco |  | Cannabis |  |
| --- | --- | --- | --- | --- | --- | --- |
| Baseline AUS/DUS assessment | CHR<br>(n = 737) | Controls<br>(n = 275) | CHR | Controls | CHR | Controls |
| Abstinent, N (%) | 444<br>(60.24%) | 134<br>(48.73%) | 562<br>(76.26%) | 255<br>(92.73%) | 564<br>(76.53%) | 246<br>(89.45%) |
| Use without impairment | 272<br>(36.91%) | 141<br>(51.27%) | 163<br>(22.12%) | 20 (7.27%) | 145<br>(19.67%) | 28 (10.18%) |
| Abuse | 17 (2.31%) | 0(0%) | 5 (.68%) | 0(0%) | 26 (3.53%) | 0(0%) |
| Dependence | 4(.54%) | 0(0%) | 7 (.95%) | 0(0%) | 2 (.27%) | 0(0%) |

| NAPLS 3 | Alcohol |  | Tobacco |  | Cannabis |  |
| --- | --- | --- | --- | --- | --- | --- |
| Baseline AUS/DUS assessment | CHR<br>(n = 551) | Controls<br>(n = 96) | CHR | Controls | CHR | Controls |
| Abstinent, N (%) | 344<br>(62.43%) | 52 (53.17) | 40<br>(7.26) | 90<br>(93.75) | 404 (73.32) | 85<br>(88.54) |
| Use without impairment | 191<br>(34.66) | 43 (44.79) | 99 (17.97) | 6 (6.25) | 110 (19.96) | 11 (11.46) |
| Abuse | 14<br>(2.54) | 1 (1.04) | 6 (1.09) | 0 (0.0) | 29 (5.26) | 0 (0.0) |
| Dependence | 2(.36%) | 0 (0.0) | 6 (1.09) | 0 (0.0) | 7 (1.27) | 0 (0.0) |

No 22qDel participants endorsed substance abuse or dependence in the past 6 months at baseline or follow-up timepoints. 22qDel carriers endorsed significantly lower rates of alcohol use than controls at both baseline (*b =* -.217, *q =* .007) and longitudinally (*b =* -.336, *q =* .007), and lower rates of non-prescription drug use at one-year follow-up (*b =* -.284, *q =* .007). Less than 4% of 22qDel carriers endorsed substance use at any timepoint, compared to 20% of TD controls endorsing substance use (See Table 2 for full results).

### Relationships between psychosis symptom domains and substance use

Due to the lack of substance use in 22qDel participants, relationships between substance use and clinical symptoms could only be investigated in CHR-P (NAPLS 2 and NAPLS 3) participants (Tables 5 and S3). In both cohorts of CHR-P participants, positive symptoms were significantly positively associated only with cannabis use severity and frequency, and baseline positive symptom severity was positively associated with cannabis use severity but not frequency at follow-up in CHR-P participants (Table 5).

**Table 5.** Results of cross-sectional models in NAPLS 2

| **Cross-Sectional** | Effect of Substance Use: Control Subjects |  | Effect of Substance Use: CHR-P Subjects |  |
| --- | --- | --- | --- | --- |
| **Positive Symptoms** | $\beta$ | $q$-value | $\beta$ | $q$-value |
| Alcohol Use Frequency | .041 | .615 | .044 | .126 |
| Alcohol Use Severity | -.015 | .739 | .018 | .299 |
| Cannabis Use Frequency | .129 | .169 | **.096** | **.011** |
| Cannabis Use Severity | .113 | .187 | **.105** | **.010** |
| Tobacco Use Frequency | .106 | .226 | .026 | .251 |
| Tobacco Use Severity | .052 | .583 | .064 | .067 |
| **Dysphoric Mood** |  |  |  |  |
| Alcohol Use Frequency | .074 | .071 | .028 | .065 |
| Alcohol Use Severity | .046 | .133 | **.068** | **.009** |
| Cannabis Use Frequency | **.211** | **<.001** | **.071** | **.009** |
| Cannabis Use Severity | **.125** | **.030** | **.083** | **.006** |
| Tobacco Use Frequency | **.112** | **.035** | **.073** | **.009** |
| Tobacco Use Severity | .060 | .107 | **.067** | **.011** |
| **Social Anhedonia** |  |  |  |  |
| Alcohol Use Frequency | -.075 | .324 | **-.139** | **<.001** |
| Alcohol Use Severity | -.051 | .324 | **-.121** | **<.001** |
| Cannabis Use Frequency | -.055 | .324 | **-.092** | **.003** |
| Cannabis Use Severity | -.056 | .324 | **-.116** | **<.001** |
| Tobacco Use Frequency | .014 | .521 | **-.127** | **<.001** |
| Tobacco Use Severity | .005 | .552 | **-.164** | **<.001** |
| **IQ** |  |  |  |  |
| Alcohol Use Frequency | .131 | .080 | **.199** | **<.001** |
| Alcohol Use Severity | .109 | .127 | **.169** | **<.001** |
| Cannabis Use Frequency | -.030 | .648 | **.086** | **.029** |
| Cannabis Use Severity | .033 | .627 | **.102** | **.010** |
| Tobacco Use Frequency | -.018 | .689 | .016 | .427 |
| Tobacco Use Severity | .027 | .653 | .021 | .393 |
| **GFS highest** |  |  |  |  |
| Alcohol Use Frequency | **.187** | **.014** | **.178** | **<.001** |
| Alcohol Use Severity | **.199** | **.013** | **.173** | **<.001** |
| Cannabis Use Frequency | .026 | .603 | .035 | .106 |
| Cannabis Use Severity | .049 | .487 | **.066** | **.026** |
| Tobacco Use Frequency | -.040 | .545 | **.097** | **.004** |
| Tobacco Use Severity | -.019 | .637 | **.125** | **<.001** |

**Table 6.** Relationships between baseline clinical measures and substance use at one-year follow-up in NAPLS-2 CHR-P

| **Positive Symptoms** | $\beta$ | *q*-value |
| --- | --- | --- |
| Alcohol Use Frequency | .007 | .872 |
| Alcohol Use Severity | .016 | .715 |
| Cannabis Use Frequency | .054 | .262 |
| Cannabis Use Severity | **.095** | **.046** |
| Tobacco Use Frequency | .007 | .889 |
| Tobacco Use Severity | -.009 | .847 |
| **IQ** |  |  |
| Alcohol Use Frequency | **.178** | **<.001** |
| Alcohol Use Severity | **.158** | **<.001** |
| Cannabis Use Frequency | **.118** | **.019** |
| Cannabis Use Severity | **.133** | **.008** |
| Tobacco Use Frequency | -.006 | .902 |
| Tobacco Use Severity | 0 | .995 |
| **Dysphoric Mood** |  |  |
| Alcohol Use Frequency | 0 | .998 |
| Alcohol Use Severity | 0 | .985 |
| Cannabis Use Frequency | 0 | .996 |
| Cannabis Use Severity | .027 | .579 |
| Tobacco Use Frequency | **.120** | **.012** |
| Tobacco Use Severity | **.113** | **.019** |
| **Social Anhedonia** |  |  |
| Alcohol Use Frequency | **-.166** | **<.001** |
| Alcohol Use Severity | **-.152** | **<.001** |
| Cannabis Use Frequency | **-.172** | **<.001** |
| Cannabis Use Severity | **-.185** | **<.001** |
| Tobacco Use Frequency | **-.221** | **<.001** |
| Tobacco Use Severity | **-.193** | **<.001** |
| **Anxiety Diagnosis** |  |  |
| Alcohol Use Frequency | -.009 | .843 |
| Alcohol Use Severity | -.033 | .486 |
| Cannabis Use Frequency | -.027 | .591 |
| Cannabis Use Severity | -.032 | .521 |
| Tobacco Use Frequency | -.009 | .856 |
| Tobacco Use Severity | .004 | .940 |
| **GFS past year highest** |  |  |
| Alcohol Use Frequency | **.145** | **<.001** |
| Alcohol Use Severity | **.109** | **.009** |
| Cannabis Use Frequency | .028 | .241 |
| Cannabis Use Severity | .032 | .222 |
| Tobacco Use Frequency | **.099** | **.021** |
| Tobacco Use Severity | **.102** | **.020** |

Dysphoric mood at baseline in CHR-P in NAPLS 2 was positively associated with cannabis use frequency and severity, tobacco use frequency and severity, and alcohol use severity (Table 5). In NAPLS 2, social anhedonia was inversely associated with substance use across all domains in CHR-P at baseline. Further, social anhedonia at baseline in CHR-P was inversely associated with substance use across all domains at one-year follow up. (Table 5). Baseline dysphoric mood in CHR-P in NAPLS 2 was positively associated with tobacco use frequency but no other substance use domain at follow-up. In NAPLS 3, dysphoric mood was positively associated with tobacco use frequency and severity in CHR-P and social anhedonia was inversely associated with substance use across all domains in CHR-P at baseline.

### Relationship between social functioning and substance use

In NAPLS 2, alcohol use frequency and severity were both positively associated with social functioning in controls at baseline. In CHR-P, baseline social functioning was positively associated with substance use across all domains at baseline and was positively associated with all substance use domains at follow-up (Table 5). While the social functioning results in CHR-P replicated in NAPLS 3, alcohol use was not associated with social functioning in controls in NAPLS 3 (Table S3).

### 22qDel carriers vs. CHR-P: group differences in neurobehavioral symptoms

The CHR-P cohort (NAPLS 2) exhibited increased positive symptom severity, dysphoric mood and social anhedonia, as well as higher IQ scores compared to 22qDel carriers, both at baseline and longitudinally (Table 7). In contrast, 22qDel carriers showed lower global social functioning than CHR-P, both at baseline and longitudinally. 22qDel carriers exhibited significantly higher rates of autism spectrum disorder compared to CHR-P.

**Table 7.**
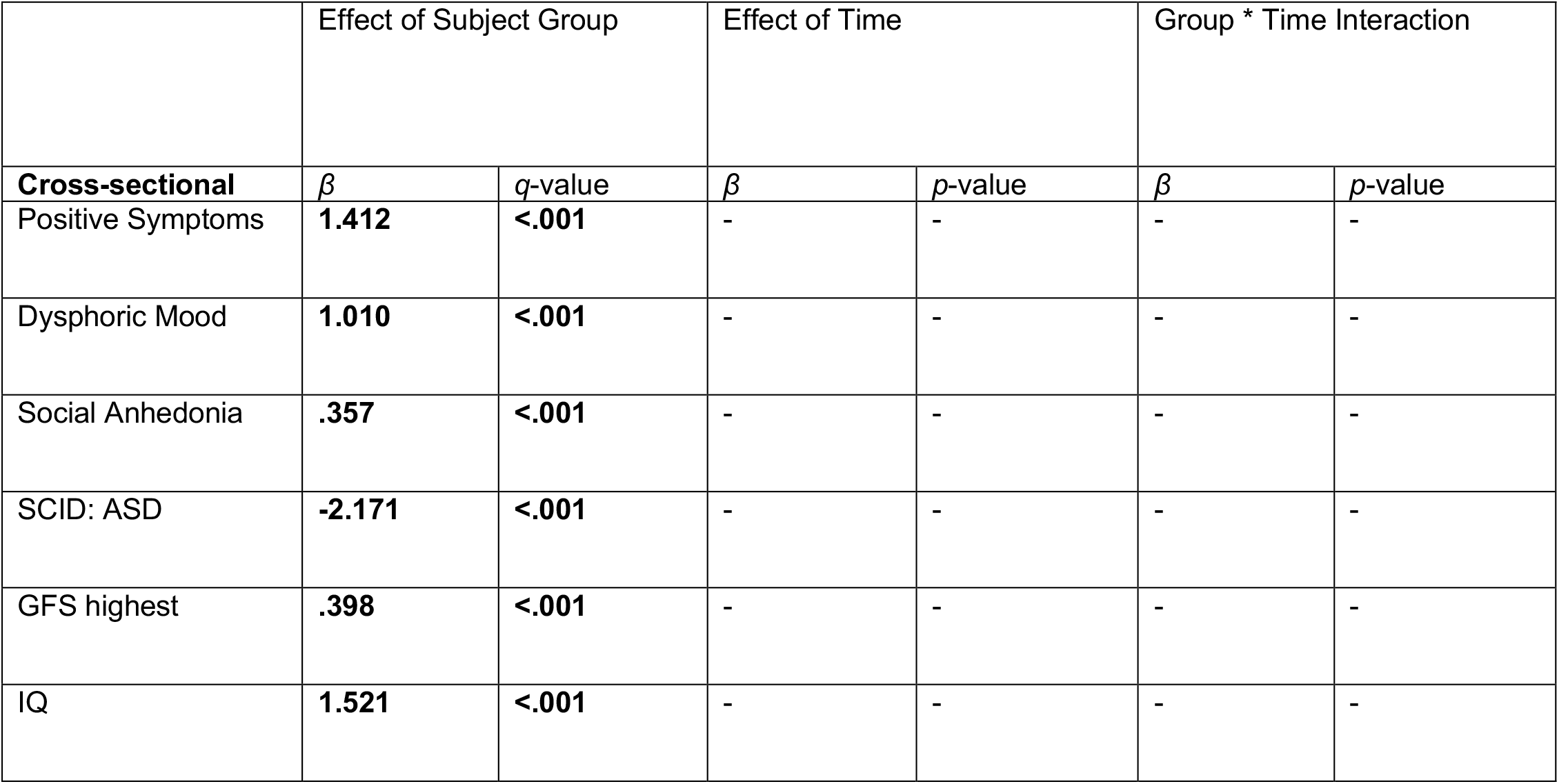
Differences in neurobehavioral measures between 22qDel and NAPLS CHR subject groups

**S2.** Differences in Substance Use between CHR-P and TD control Groups in NAPLS 3

|  | Effect of Subject Group |  | Effect of Time |  | Group * Time Interaction |  |
| --- | --- | --- | --- | --- | --- | --- |
| **Cross-sectional** | $\beta$ | *p*-value | $\beta$ | *q*-value | $\beta$ | *p*-value |
| Alcohol Use Frequency | -.082 | .136 | - | - | - | - |
| Alcohol Use Severity | -.088 | .129 | - | - | - | - |
| Cannabis Use Frequency | **.297** | **.003** | - | - | - | - |
| Cannabis Use Severity | **.380** | **<.001** | - | - | - | - |
| Tobacco Use Frequency | **.299** | **.002** | - | - | - | - |
| Tobacco Use Severity | **.343** | **<.001** | - | - | - | - |
| GFS | **-1.416** | **<.001** | - | - | - | - |
| Positive Symptoms (SIPS) | **2.37** | **<.001** | - | - | - | - |
| Negative Symptoms | **1.543** | **<.001** | - | - | - | - |
| Disorganized Symptoms | **1.377** | **<.001** | - | - | - | - |
| General Symptoms | **1.695** | **<.001** | - | - | - | - |
| Dysphoric Mood | **1.720** | **<.001** | - | - | - | - |
| Social Anhedonia | **1.216** | **<.001** | - | - | - | - |

**S3.**
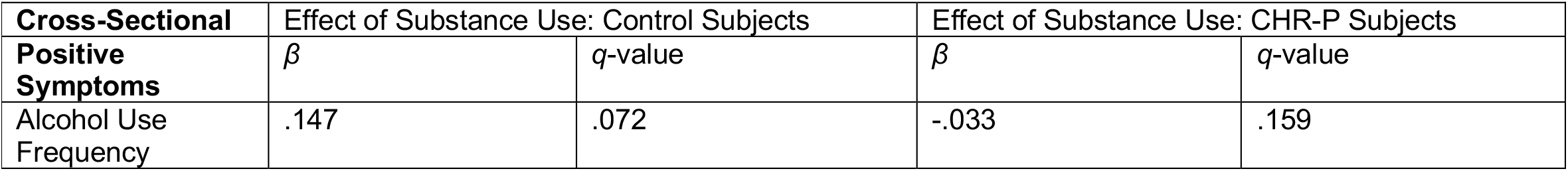
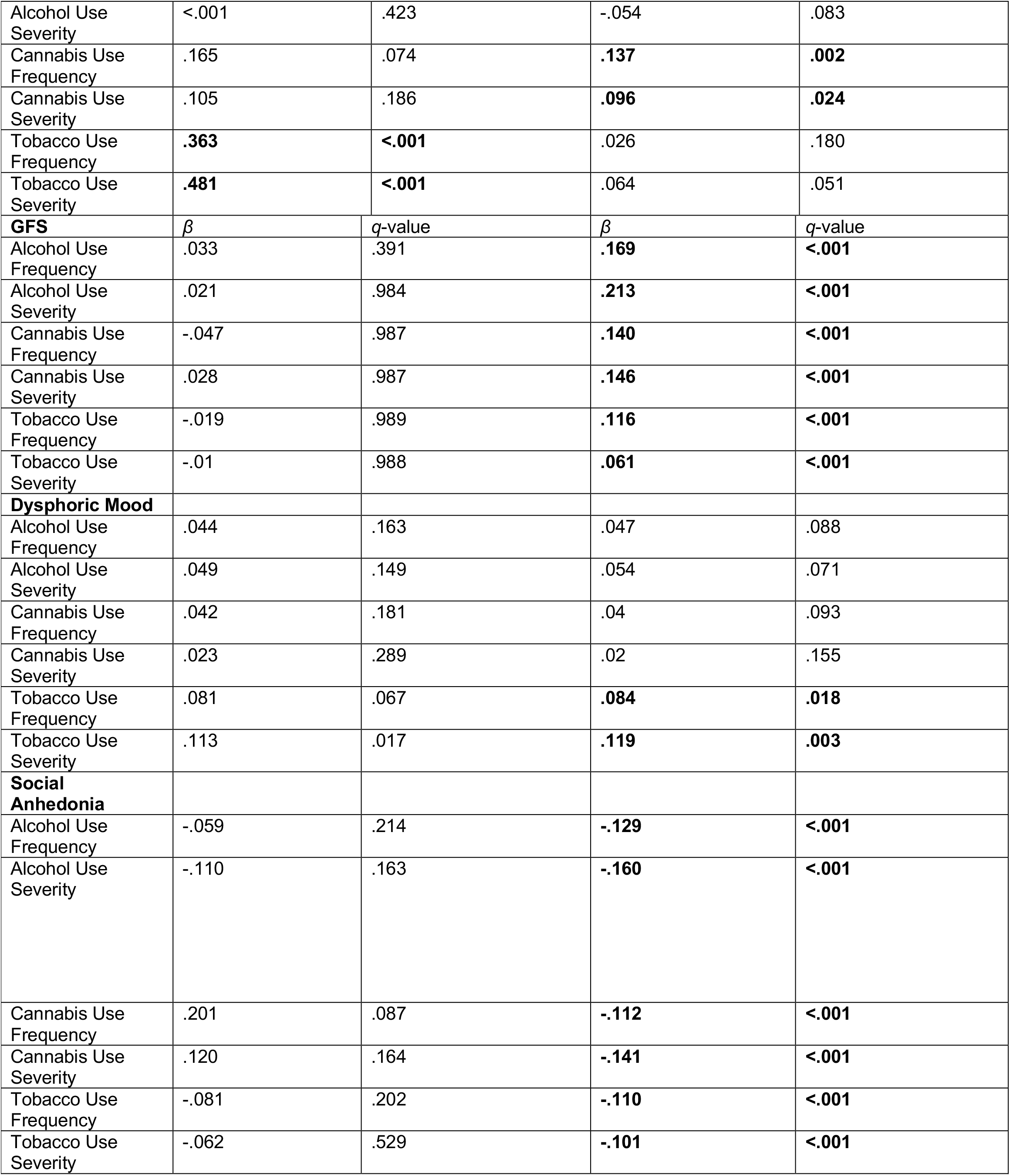
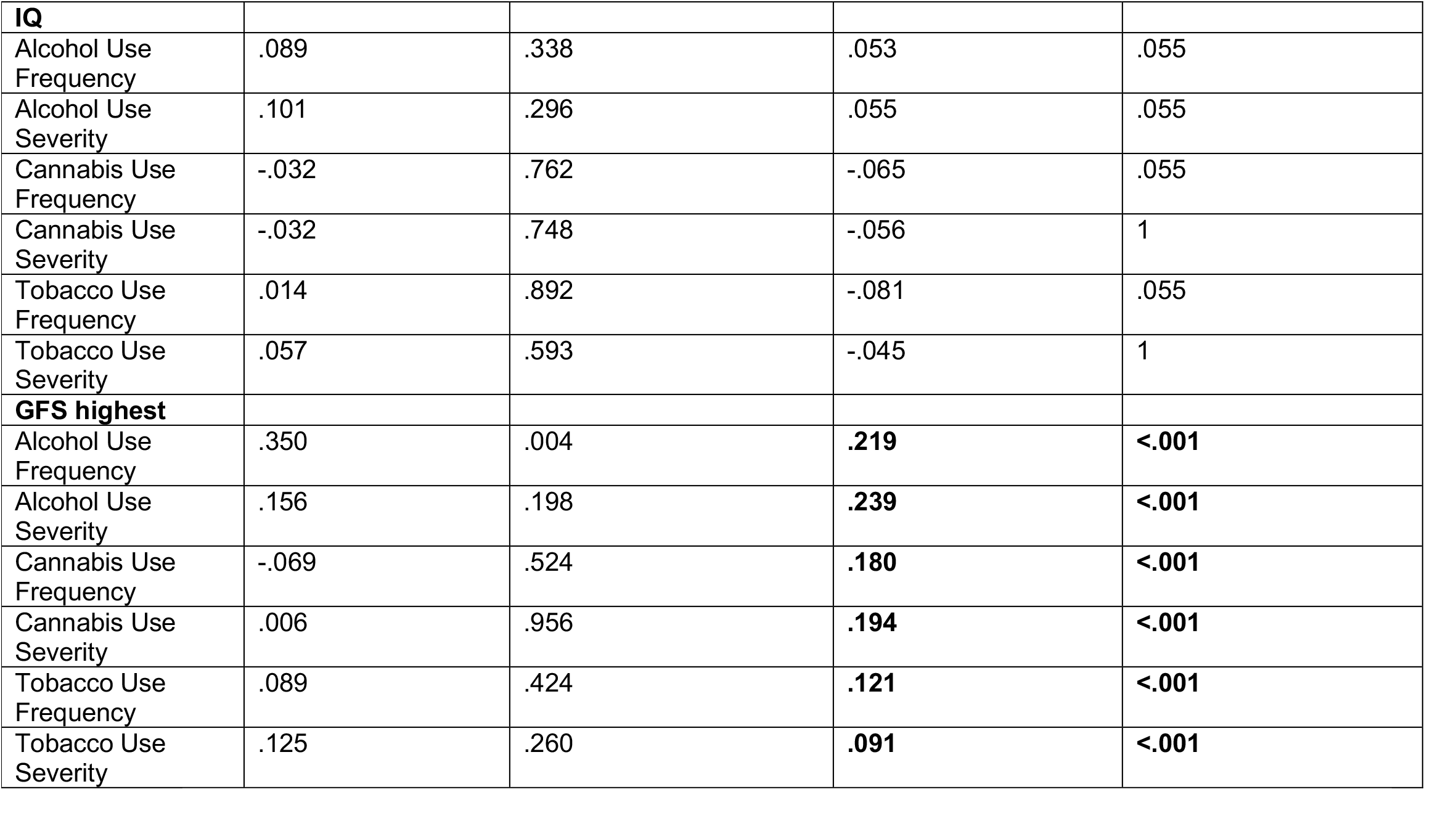
NAPLS 3 Relationships between neurobehavioral measures and substance use in TD controls and CHR-P

| **Cross-Sectional** | Effect of Substance Use: Control Subjects |  | Effect of Substance Use: CHR-P Subjects |  |
| --- | --- | --- | --- | --- |
| **Positive Symptoms** | $\beta$ | *q*-value | $\beta$ | *q*-value |
| Alcohol Use Frequency | .147 | .072 | -.033 | .159 |
| Alcohol Use Severity | <.001 | .423 | -.054 | .083 |
| Cannabis Use Frequency | .165 | .074 | **.137** | **.002** |
| Cannabis Use Severity | .105 | .186 | **.096** | **.024** |
| Tobacco Use Frequency | **.363** | **<.001** | .026 | .180 |
| Tobacco Use Severity | **.481** | **<.001** | .064 | .051 |
| **GFS** | $\beta$ | $q$-value | $\beta$ | $q$-value |
| Alcohol Use Frequency | .033 | .391 | **.169** | **<.001** |
| Alcohol Use Severity | .021 | .984 | **.213** | **<.001** |
| Cannabis Use Frequency | -.047 | .987 | **.140** | **<.001** |
| Cannabis Use Severity | .028 | .987 | **.146** | **<.001** |
| Tobacco Use Frequency | -.019 | .989 | **.116** | **<.001** |
| Tobacco Use Severity | -.01 | .988 | **.061** | **<.001** |
| **Dysphoric Mood** |  |  |  |  |
| Alcohol Use Frequency | .044 | .163 | .047 | .088 |
| Alcohol Use Severity | .049 | .149 | .054 | .071 |
| Cannabis Use Frequency | .042 | .181 | .04 | .093 |
| Cannabis Use Severity | .023 | .289 | .02 | .155 |
| Tobacco Use Frequency | .081 | .067 | **.084** | **.018** |
| Tobacco Use Severity | .113 | .017 | **.119** | **.003** |
| **Social Anhedonia** |  |  |  |  |
| Alcohol Use Frequency | -.059 | .214 | **-.129** | **<.001** |
| Alcohol Use Severity | -.110 | .163 | **-.160** | **<.001** |
| Cannabis Use Frequency | .201 | .087 | **-.112** | **<.001** |
| Cannabis Use Severity | .120 | .164 | **-.141** | **<.001** |
| Tobacco Use Frequency | -.081 | .202 | **-.110** | **<.001** |
| Tobacco Use Severity | -.062 | .529 | **-.101** | **<.001** |
| **IQ** |  |  |  |  |
| Alcohol Use Frequency | .089 | .338 | .053 | .055 |
| Alcohol Use Severity | .101 | .296 | .055 | .055 |
| Cannabis Use Frequency | -.032 | .762 | -.065 | .055 |
| Cannabis Use Severity | -.032 | .748 | -.056 | 1 |
| Tobacco Use Frequency | .014 | .892 | -.081 | .055 |
| Tobacco Use Severity | .057 | .593 | -.045 | 1 |
| **GFS highest** |  |  |  |  |
| Alcohol Use Frequency | .350 | .004 | **.219** | **<.001** |
| Alcohol Use Severity | .156 | .198 | **.239** | **<.001** |
| Cannabis Use Frequency | -.069 | .524 | **.180** | **<.001** |
| Cannabis Use Severity | .006 | .956 | **.194** | **<.001** |
| Tobacco Use Frequency | .089 | .424 | **.121** | **<.001** |
| Tobacco Use Severity | .125 | .260 | **.091** | **<.001** |

## Discussion

Our study represents, to our knowledge, the first direct comparison of substance use patterns and neurobehavioral correlates in youth at clinical and genetic high risk for psychosis, Specifically, we compared youth with 22q11.2 deletions (22qDel) to a clinically/behaviorally defined high-risk cohort (CHR-P youth), and found support for extremely low rates of substance use in 22qDel, but elevated rates of substance use in CHR-P youth, relative to TD controls. These results suggest that despite conferring elevated risk for psychosis (Olsen et al., 2018), the 22q11.2 deletion appears to be protective against initiating and/or continuing substance use. We then tested cross-sectional and longitudinal associations between substance use and neurobehavioral traits in CHR-P youth and found broadly that increased severity of positive psychosis symptoms and dysphoric mood, as well as better social functioning and higher IQ, were associated with greater substance use frequency and severity. In contrast, greater social anhedonia was associated with significantly lower substance use, across domains. These patterns persisted when we investigated these relationships longitudinally over one-year. Finally, we directly compared these neurobehavioral measures in 22qDel carriers and CHR-P youth and found that CHR-P youth exhibited significantly increased positive psychosis symptoms, dysphoric mood, social functioning, and IQ compared to 22qDel carriers, but significantly lower current social anhedonia as well as lower rates of ASD compared to 22qDel carriers.

This work expands upon prior studies reporting elevated rates of substance use in individuals at high risk for psychosis (Santesteban-Echarri et al., 2022; Farris et al., 2020; Buchy et al. 2015; Addington et al., 2014) by examining associated neurobehavioral factors both cross-sectionally and longitudinally and in parallel with a population at genetically high risk for psychosis. Substances of misuse directly or indirectly activate the mesolimbic dopamine pathway, which is associated with the reward properties of drugs and positive symptoms of schizophrenia (Volkow et al. 2009). An array of dopaminergic abnormalities have been reported in CHR-P populations, including alterations in midbrain and striatal responses to reward assessed via functional neuroimaging studies, and elevated dopamine synthesis, storage, and release, observed in positron emission tomography and single photon emission computed tomography studies (for a review see Howes et al. 2022). In addition, CHR-P individuals display impaired reward processing associated with abnormal striatal activity during task performance (Karcher et al., 2016; Walz et al., 2015), related to symptom severity. Research on 22qDel mesolimbic differences using neuroendocrine and peripheral dopaminergic markers has revealed dopaminergic abnormalities (Boot et al. 2008), along with deficits in pleasure responses and reduced activation in medial frontal areas during reward anticipation (van Duin et al. 2016; Dubourg et al. 2016). Our study supports a body of work indicating that elevated rates of cannabis use in individuals at high risk for psychosis are related to positive psychosis symptom severity, which may be mediated by alterations in these reward-related pathways (for reviews, see Farris et al., 2020, and Gunasekera et al., 2022), while there is mixed evidence on the association between alcohol or tobacco use and positive psychosis symptoms (Addington et al., 2014). By contrast, positive symptom severity was significantly lower in 22qDel carriers.

CHR-P youth displayed significantly better social functioning and reduced rates of ASD compared to 22qDel carriers, and social functioning was positively associated with substance use, both at baseline and longitudinally, within CHR-P. These results are consistent with previous descriptions of withdrawn behavior and problems with peer social interaction in 22qDel carriers (Swillen & McDonald-McGinn 2015), as well as with a previous single-site study finding higher levels of social functioning associated with elevated substance use rates in CHR-P (Author et al., 2012). CHR-P individuals report social engagement as a primary reason for substance use (Gill et al., 2015; Gomez-Perez et al., 2017; Kolliakou et al., 2015). It has been hypothesized that social skills facilitate drug acquisition in individuals with psychotic disorders such that poorer social skills make illicit substances more difficult to obtain (Mueser et al., 2000; Compton et al. 2004), especially where there are legal barriers (Rais et al. 2008). Higher peer engagement has been found to be associated with increased risk for substance use particularly during adolescence (Keefe 1994; Kuther & Higgins-D’Alessandro 2003; Whitesell et al., 2013; Brooks-Russel et al. 2014). In addition to high rates of ASD (Lin et al., 2019), social phobia is overrepresented in 22qDel (Dekker & Koot 2003) and may represent a barrier to drug acquisition and use. That social functioning is significantly lower in 22qDel carriers compared to CHR-P youth may in fact represent a protective factor against hazardous substance use for 22qDel carriers.

In both NAPLS 2 and NAPLS 3 CHR cohorts, dysphoric mood was positively associated with all substance use domains at baseline and predicted elevated tobacco use rates at follow up. This is consistent with previous reports of CHR-P individuals citing mood enhancement as motivation for seeking cannabis (Gill et al., 2015; Green 2004; Spencer 2002). That 22qDel carriers had significantly lower dysphoric mood compared to CHR-P youth may indicate that while mood enhancement is a primary motivation for drug-seeking in CHR-P individuals, this motive may not be as strong for 22qDel carriers.

Social anhedonia in CHR-P was significantly inversely correlated with substance use at baseline and predicted decreased substance use at follow-up. This supports previous findings that CHR-P cannabis users have significantly lower levels of social anhedonia compared to CHR-P non-users, proposed to be explained by lower social anhedonia driving peer engagement and substance-seeking for social motives (Gill et al., 2015; Kolliakou et al., 2015; Santesteban-Echarri et al., 2022). That levels of social anhedonia were lower in 22qDel carriers compared to CHR-P may indicate that lower rates of social anhedonia in 22qDel may not be driving peer engagement and substance use in the same way as for CHR-P individuals. Lower social functioning and elevated rates of ASD in 22qDel carriers may again serve as a protective barrier to facilitating peer engagement and substance use and acquisition. While a relationship between psychosis symptoms, specifically increased grandiosity and lowered social anhedonia, and cannabis use have been reported in NAPLS 3 (Santesteban-Echarri et al., 2022), our work expands upon these findings by focusing specifically on the enhanced sample of participants and investigating alcohol and tobacco use. Results broadly replicated in our enhanced CHR-P sample (NAPLS 3), providing support that these findings hold in a population at especially high risk for psychosis.

Consistent with previous literature (Ferraro et al., 2019), here we also found that the likelihood of substance use increased with higher IQ within the CHR-P group. IQ scores in CHR-P youth were, on average, significantly higher compared to those of 22qDel carriers. While we could not test the relationship between IQ and substance use in 22qDel patients given the minimal substance use reported in our cohort, a previous study reported substance use increased with higher IQ scores in 22qDel carriers (Vingerhoets et al., 2019). This prior work speculated as an explanation for these findings that individuals with more severe intellectual disability (ID) are more likely to be cared for or protected by their family members or institutions providing supervision. Substance use rates are relatively low in individuals with idiopathic ID compared to TD individuals, similarly to 22qDel; however, compared to individuals with substance use disorders (SUD) without ID, individuals with ID and SUD are less likely to receive treatment or to remain in treatment (Chapman & Wu, 2012; van Duijvenbode et al., 2015). Some variables found to influence substance use in individuals with ID include social pressure and the desire to increase social inclusion and overcome loneliness (Christian. & Poling, 1997; Degenhardt, 2000; Kalyva, 2007). Future work should investigate whether 22qDel carriers who do have hazardous substance use patterns are undertreated.

The current study has several important clinical implications. That 22qDel, despite conferring increased genetic risk for psychosis, is protective against problematic substance use offers insight into behavioral risk factors that can be targeted for intervention in individuals at CHR-P. Drugs of abuse can contribute to the positive symptoms of psychosis, and people with any substance use disorder have an earlier age of schizophrenia onset (for a review, see Hunt et al. 2018). Cannabis in particular can be a contributory risk factor for psychotic disorder, and animal models show that the developing brain is susceptible to cannabis-induced brain morphological and circuitry changes (Hall & Degenhardt 2008; Realini et al. 2011). As cannabis legalization becomes more prevalent, it is important to identify avenues for behavioral intervention for CHR-P as access increases. Our findings suggest the importance of neurobehavioral risk factors and social context in influencing substance use in CHR-P, and abstinence in 22qDel, pointing to an intervention that targets social influence in disrupting the initiation and continuation of substance use. Strengths of this study include the large sample of individuals at CHR-P and the sample of participants with 22qDel, a relatively rare disorder. Adolescence and young adulthood are crucial developmental periods for studying the relationship between substance use and psychosis (e.g. Henquet et al. 2008); our study provides longitudinal insight into the progression of clinical symptoms and behavioral trajectories during neurodevelopment, whereas many previous studies utilize cross-sectional designs. Previous literature has focused primarily on the relationship between cannabis use and schizophrenia rather than psychosis spectrum disorders, and mixed findings have been attributed to differences in the temporal relationship between onset of cannabis use and onset of psychosis (Machielsen et al. 2010). Most previous studies on substance use in psychiatric populations lack details on the severity and frequency of use, and often conflate types of substance use, and very few include a healthy comparison group.

This study also has several important limitations. A majority of NAPLS data was collected pre-legalization of marijuana in the United States, and rates of substance use in this sample may become greater in regions where marijuana is legalized. Details on cannabis use including type of cannabis, dose, and whether cannabidiol was also consumed were not collected. Urine toxicology data would have also provided important biological information on cannabis use in the sample. Future directions include studying cannabis consumption at a finer level of detail. In NAPLS 2 there were few “abuse” and “dependence” occurrences recorded for substance use, and this may limit an ability to detect a contribution of heavy substance use to psychotic transition and/or clinical outcome. Demands of the NAPLS studies may also have deterred heavier substance users from participating, which may have biased our samples. While psychotropic medication usage rates are comparable amongst CHR-P and 22qDel patients and controlling for medication use did not affect our results, other medical issues specific to 22qDel may also explain decreased substance use. Extremely low rates of substance use in 22qDel carriers rendered this study underpowered to detect associations between substance use and neurobehavioral traits in 22qDel carriers.

In conclusion, we found that despite higher rates of substance use in CHR-P individuals compared with controls, individuals with 22qDel, although at increased risk for psychosis, had markedly lower prevalence of substance use relative to TD controls. As this is the first study to investigate neurobehavioral traits underlying substance use patterns in CHR-P compared to 22qDel carriers, more research is needed into other factors such as environmental risk that may interplay with genetic factors to confer this effect. 22qDel could be a valuable model to study genetic factors in addition to behavioral factors underlying substance use in the general population.

## Data Availability

All data produced in the present study are available upon reasonable request to the authors

